# Contribution of moderation foods to total energy intake by sociodemographic characteristics in U.S. adults: National Health and Nutrition Examination Survey, 2017 – 2020

**DOI:** 10.1101/2025.09.30.25336983

**Authors:** Leah M. Lipsky, Amara J. Channell Doig, Tonja R. Nansel

## Abstract

**Background:** U.S. adults exceed recommended intakes of multiple nutrients of concern

**Objective:** This study uses a recently-developed food-level classification method for identifying foods exceeding thresholds for nutrients of concern to investigate sociodemographic differences in their contribution to total energy intake among U.S. adults.

**Design:** Cross-sectional.

**Participants/setting:** Participants 20 – 80 years of age in the 2017–2020 National Health and Nutrition Examination Survey (n=7706).

**Main outcome measures:** Data from 24-hour dietary recalls were used to identify “moderation foods” meeting at least one threshold for high content of nutrients of concern: added sugar >20% energy, sodium >460mg per serving, refined grains >50% of total grains or >10:1 ratio of carbohydrate to fiber content, saturated fat >20% energy, total fat >9% by weight (applied only to vegetables, sweets, and snacks), and all alcoholic beverages. The percentage of energy intake from moderation foods in total and from foods meeting each of the moderation food thresholds were calculated and compared across age, education, income-to-poverty ratio (IPR), and race/ethnicity.

**Statistical analyses performed:** Linear regression models with multiplicative interaction terms tested whether race/ethnicity or education modified relationships of IPR with moderation food intake.

**Results:** Moderation foods contributed 76% (95%CI:75%-77%) of energy intake overall. Intake was higher in younger adults (mean=79%) versus older (72%-75%) adults, males (77%) versus females (74%), and adults in the lowest (79%) vs. highest (74%) income quartile. Moderation food intake was lowest in Non-Hispanic Asian adults (66%) and among adults with the highest education (mean=71%). Foods meeting thresholds for sodium, refined grains, and fat contributed the most to overall energy intake. Foods meeting thresholds for added sugar, sodium, and refined grains contributed a smaller share of energy intake in non-Hispanic Asians versus other racial/ethnic groups and in older versus younger adults. Income was inversely associated with moderation food intake only among high school graduates (second highest education group; β = - 1.1 95%CI = -2.1 to -0.2) and modestly inversely associated with moderation food intake in all race/ethnic groups except Mexican Americans/Other Hispanic adults.

**Conclusions:** High intake of moderation foods across all subgroups of U.S. adults, with only modest between-group differences in the contributions of different categories of moderation foods, suggest a need for population-wide changes in food selection to align with dietary guidelines. In particular, interventions and policies to assist in identifying foods high in sodium, refined grains, and fat may help consumers improve food choices.

## Introduction

The 2020-2025 Dietary Guidelines for Americans (DGA) recommend limiting intake of added sugar, saturated fat, sodium, refined grains, and alcoholic beverages (“moderation components”) due to their associations with multiple adverse health outcomes and their tendency to displace intake of foods that provide essential nutrients^1,2^. However, most Americans exceed recommended maximum intakes of refined grains (>90%), added sugar (70%), saturated fat (70%), and sodium (90%), contributing to the high public health burden of diet-related chronic diseases^1^.

While sociodemographic differences in overall diet quality have been reported in multiple studies, these differences are generally modest in magnitude. For example, 2018 – 2018 NHANES data indicate relatively small mean differences (statistical comparisons not provided) in Healthy Eating Index-2015 scores (an indicator of conformance to the 2015-2020 Dietary Guidelines for Americans) between females (mean HEI=60) and males (mean HEI=56), non-Hispanic Asian (mean HEI=65) and Hispanic (mean HEI=58) adults versus non-Hispanic white (mean HEI=57) and non-Hispanic black (mean HEI=54) adults, and in adults with household income greater than 350% of the federal poverty level (mean HEI=60) versus those with incomes less than 350% of the federal poverty level (mean HEI=56)^3^. Education is also positively associated with better diet quality^4–8^, and some evidence suggests that associations of income with diet quality differ by education and race/ethnicity^8,9^. In the U.S., educational attainment is positively correlated with income^10^, and both income and education differ by race/ethnicity^11,12^. However, few previous studies on sociodemographic correlates of diet quality have examined the potential for confounding or interactions between these variables.

Additionally, research on sociodemographic differences in intake of moderation dietary components has focused on individual nutrients or food groups (such as sugar-sweetened beverages or snack foods)^13–16^, ultra-processed foods (based on processing rather than nutritional components)^4,17,18^, or “junk”^19^ or “fast”^20^ foods (using multiple definitions and typically applied only to select food groups). Given the lack of a guidelines-based definition that can be applied to all foods in the Food and Nutrient Database for Dietary Studies (FNDDS), we previously developed a method to identify moderation foods based on evidence-based thresholds for added sugar, sodium, fat, refined grains, and alcohol. Although some moderation foods may also provide beneficial nutrients, initial validation analyses indicated that the method effectively captures foods that are disproportionately high in nutrients of concern (by distinguishing between foods with low versus high nutrient density) and that greater intake of moderation foods is associated with lower overall adherence to dietary guidelines in U.S. adults (r = -0.72, p<0.0001 with Healthy Eating Index-2020 scores)^21^.

Examining the distribution of moderation food intake across population subgroups using a systematic, guidelines-based indictor will help clarify how moderation food contributes to socioeconomic differences in overall diet quality and identify subgroups who may benefit most from targeted interventions. This study aimed to compare moderation food intake across sociodemographic characteristics among U.S. adults and investigate whether education or race/ethnicity modifies associations of income with moderation food intake.

## Methods

### Study design and population

This study used publicly available data from the pre-pandemic cycle (2017 – 2020) of the National Health and Nutrition Examination Survey (NHANES), administered by the National Center for Health Statistics (NCHS). NHANES is a population-based, cross-sectional survey that uses a complex, stratified, multistage probability sample design to generate a representative sample of the civilian, non-institutionalized U.S. population. Details on the study design, protocol, and data collection have been documented elsewhere^22^. The protocols are approved by the NCHS Research Ethics Review Board, and informed consent was obtained from participants ≥ 18 years of age.

The analytic sample for this study included adults at least 20 years old with at least one 24-hour dietary recall determined to be reliable by NCHS^23^, and data on at least one of the sociodemographic variables. Of the n = 9332 total adult participants, n = 7706 were included in the final analytic data set. This secondary analysis of deidentified publicly available data was exempt from institutional review board review.

### Data collection

Participants self-reported sociodemographic characteristics during an in-home interview administered by trained personnel. Dietary intake data come from the What We Eat In America dietary interview component of NHANES; trained interviewers conducted two 24-hour dietary recalls using the USDA’s Automated Multiple-Pass Method^24^, with the first recall administered in-person at the mobile exam center and the second by telephone approximately 3 – 10 days after the initial recall.

### Moderation food intake

Foods reported in the 24-hour diet recalls were classified as moderation foods if they met any of the previously established criteria, described in detail elsewhere^21^. Briefly, component thresholds included: added sugar >20% of energy; sodium >460 mg per serving; refined grains >50% of total grains (ounce equivalents) and >10:1 carbohydrate: fiber ratio (g); saturated fat >20% of energy (excluding vegetables); total fat >9% of energy (applied only to vegetables, sweets, and snacks), and all alcoholic beverages^21^. Foods that did not exceed any threshold were classified as non-moderation. Total moderation food intake (% total energy intake, kcal) was calculated from the sum of energy intake from moderation food relative to total energy intake across both diet recalls. The percent of energy intake (% kcal) from foods meeting each of the individual moderation food component thresholds was also calculated.

### Sociodemographic characteristics

NHANES categorizes age as 20 – 39 y, 40 – 59 y, and ≥ 60 y^25^. Race/ethnicity is categorized as Mexican American/Hispanic, non-Hispanic white, non-Hispanic black, non-Hispanic Asian, and multi/other. Education categories include less than high school, high school degree/general equivalency diploma, some college, college graduate or above. Income poverty ratio (IPR)^26^ is a continuous measure that reflects household income relative to the federal poverty threshold, accounting for household size and composition. NHANES truncates the maximum IPR at 5 due to disclosure concerns; IPR quartiles were used for analysis.

### Statistical analysis

All analyses used survey methods to account for the complex sampling design of NHANES using StataSE version 18. Frequencies and percentages were calculated for sociodemographic characteristics, and means and 95% confidence intervals of moderation food intake (% kcal) and intake (% kcal) from foods meeting each of the moderation food thresholds, were calculated for each sociodemographic subgroup. Differences in mean moderation food intake by sociodemographic characteristics were evaluated using linear regressions and post-estimation pairwise comparisons; Sidak adjustment was used to account for multiple comparisons. Separate models with multiplicative interaction terms were used to investigate whether education or race/ethnicity modified associations of IPR (examined as a continuous variable) with moderation food intake.

## Results

The sample was approximately evenly split according to age group and sex (**Table 1**). Sixty-three percent of the sample was non-Hispanic white. Ten percent of the weighted sample earned less than a high school degree, and the remainder was approximately evenly split between education groups. IPR was disproportionately concentrated at the highest truncated value (5), with the mean ± SD of the upper quartile = 4.9±0.1.

**Table 1.**
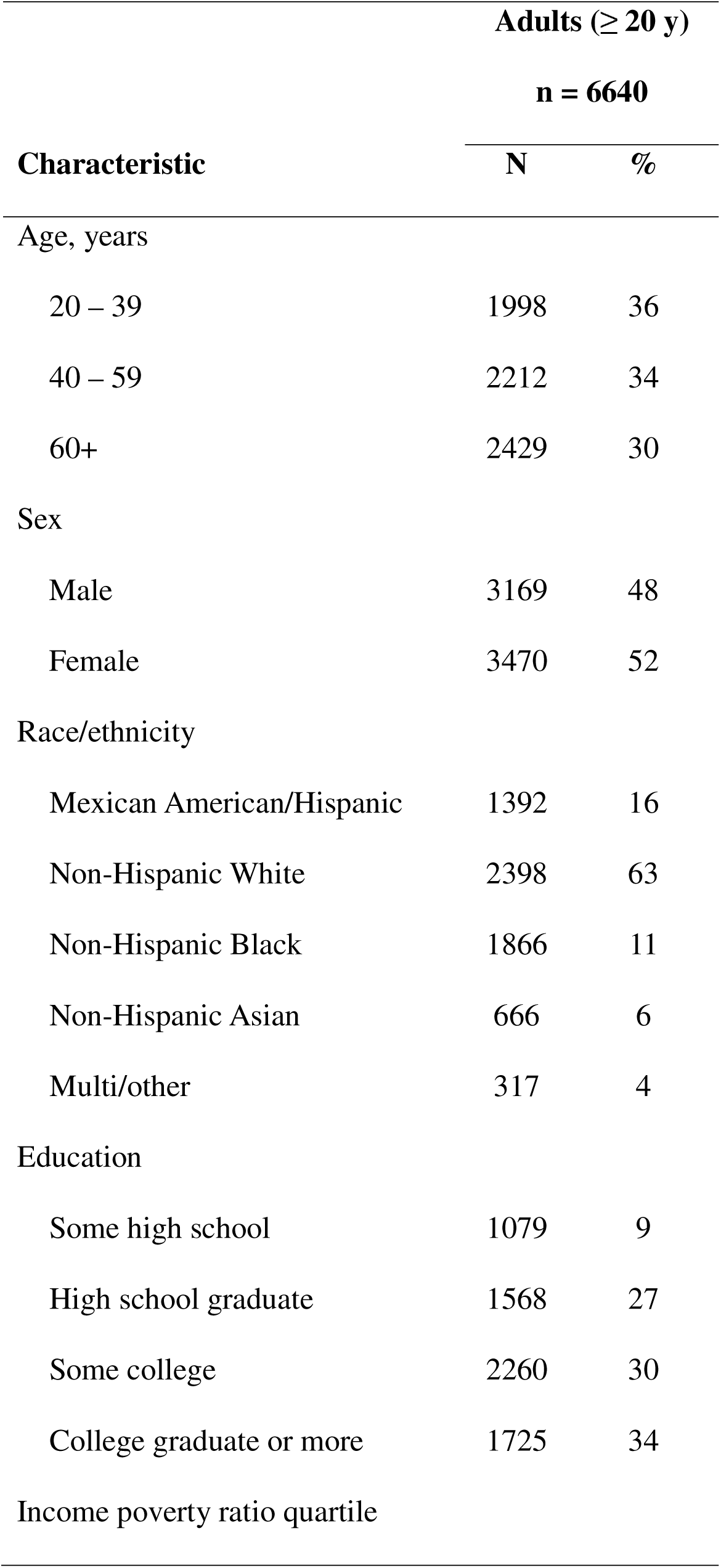
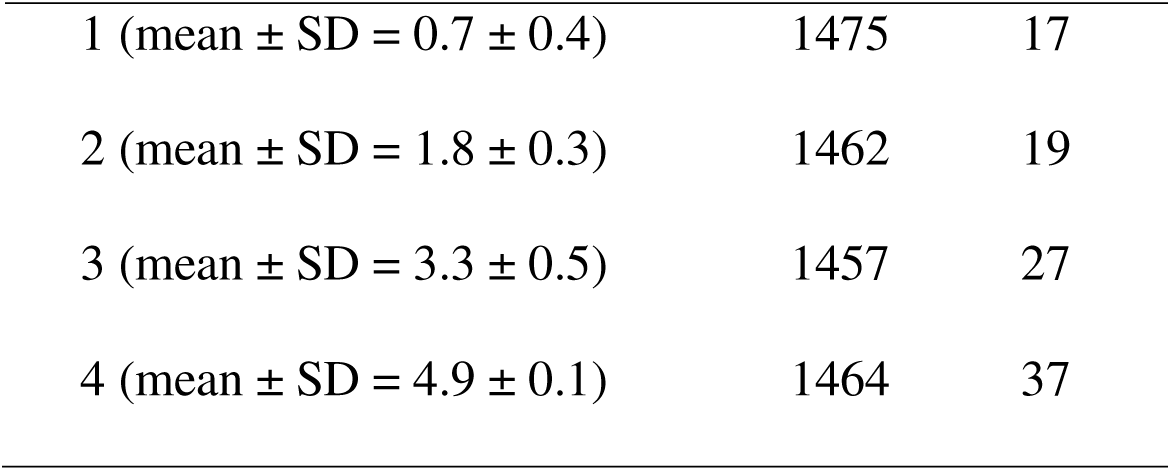
Sample characteristics.

|  | **Adults ($\geq 20$ y)** |  |
| --- | --- | --- |
|  | **n = 6640** |  |
| **Characteristic** | **N** | **%** |
| Age, years |  |  |
| 20 – 39 | 1998 | 36 |
| 40 – 59 | 2212 | 34 |
| 60+ | 2429 | 30 |
| Sex |  |  |
| Male | 3169 | 48 |
| Female | 3470 | 52 |
| Race/ethnicity |  |  |
| Mexican American/Hispanic | 1392 | 16 |
| Non-Hispanic White | 2398 | 63 |
| Non-Hispanic Black | 1866 | 11 |
| Non-Hispanic Asian | 666 | 6 |
| Multi/other | 317 | 4 |
| Education |  |  |
| Some high school | 1079 | 9 |
| High school graduate | 1568 | 27 |
| Some college | 2260 | 30 |
| College graduate or more | 1725 | 34 |
| Income poverty ratio quartile |  |  |
| 1 (mean ± SD = 0.7 ± 0.4) | 1475 | 17 |
| 2 (mean ± SD = 1.8 ± 0.3) | 1462 | 19 |
| 3 (mean ± SD = 3.3 ± 0.5) | 1457 | 27 |
| 4 (mean ± SD = 4.9 ± 0.1) | 1464 | 37 |

Mean moderation food intake (% kcal) in the full sample was 76% (95% CI = 75% - 77%). Intakes from foods meeting thresholds for sodium, refined grains, and fat were 12 to 31 percentage points higher than that from foods meeting thresholds for sugar and alcohol (**Supplemental Table 1**).

Moderation food intake by sociodemographic characteristics are presented in **Figures 1 – 5** and Supplemental Table 1. Mean moderation food intake varied from 79% (95%CI: 78% - 80%) in adults 20 – 39 years of age to 72% (95%CI: 71% - 73%) in adults ≥ 60 years of age (p < 0.001 for all pairwise comparisons) (**Figure 1a**). Mean moderation food intake was 4.1% kcal (SE = 0.7% kcal) lower in 40 – 59-year-olds versus 20 – 39-year-olds (p<0.001), 7.2% kcal (SE = 0.6% kcal) lower in those ≥ 60 years-old versus 20 – 39-year-olds (p<0.001), and 3.1% kcal (SE = 0.7% kcal) lower in those ≥ 60 years-old versus 40 – 59 years old (p = 0.01) (Figure 1a). **Figure 1b** and Supplemental Table 1 indicate that these differences were primarily attributable to differences in intakes of foods meeting thresholds for sodium (3 – 9 percentage points higher in middle aged and younger adults versus older adults) and refined grains (2 to 6 percentage points higher in middle aged and young adults versus older adults).

Mean moderation food intake was 3.2% kcal higher (SE=0.6% kcal) (p < 0.001) in males (77%, 95%CI: 76% - 78%) than females (74%, 95%CI: 73% - 75%) (**Figure 2**). These differences were primarily attributable to differences in intakes of foods meeting thresholds for sodium (four percentage points higher in males than females) and refined grains (two percentage points higher in males than females) (**Figure 2b** and Supplemental Table 1).

When comparing by race/ethnicity (**Figure 3**), moderation food intake was lowest in non-Hispanic Asian adults (66%, 95%CI: 64% - 69%) and highest in multi/other (80%, 95%CI: 77% - 82%) and non-Hispanic black adults (79%, 95%CI: 77% - 80%). Moderation food intake was 73% (95%CI: 72% - 75%) in Mexican American/Hispanic adults and 76% (95%CI: 75% - 78%) in non-Hispanic white adults. While differences in mean moderation food intake of multi/other as compared to non-Hispanic white (p=0.08) and black (p=0.998) adults were not statistically different, all other pairwise comparisons were statistically different (p<0.02). The relatively low moderation food intake in non-Hispanic Asians was due to lower intake of foods exceeding thresholds for refined grains, added sugar, sodium, and fat (**Figure 3b** and Supplemental Table 1). Mexican American/Hispanic adults also had relatively low intakes of refined grains, added sugar, and fat, but consumed a relatively higher amount of foods meeting the sodium threshold.

Moderation food intake was lower in adults with at least a college degree (71%, 95%CI: 70% - 73%) than in adults with less education (p < 0.001), but mean intake did not significantly differ across the three lower education groups (75% - 79%) (**Figure 4a**). Mean moderation food intake in adults with the highest education was 4.2% kcal (SE=0.9% kcal) lower than in those with less than a high school degree (p=0.008), 7.5% kcal (SE=1.1% kcal) lower than those with a high school degree only (p<0.001), and 6.1% kcal (SE=0.8% kcal) lower than those with some college education (p<0.001). Mean moderation food intake in adults with a high school degree was 3.3% (SE=0.9%) higher than in those with less than a high school degree (p=0.008). **Figure 4b** and Supplemental Table 1 indicate that the lower intake of moderation foods in the highest education group was due primarily to lower intake of foods meeting the refined grains threshold (31% versus 35% - 38% kcal). Group mean intakes from foods meeting other component thresholds were within four percentage points.

Mean moderation food intake was inversely associated with IPR quartile (**Figure 5a**), with a 4.6-point difference between the 1^st^ (79%, 95%CI: 77% - 80%) and 4^th^ quartile (74%, 95%CI: 72% - 76%) (p=0.005. However, only the pairwise differences between the 1^st^ and 3^rd^ quartiles (76%, 95%CI: 74% - 78%) and 1^st^ and 4^th^ quartiles were statistically significant (p = 0.02 and 0.005, respectively). These differences were primarily due to the relatively lower intake in the highest IPR quartile of foods meeting the added sugar threshold (17% versus 20% for the 2^nd^ and 3^rd^ quartile and 22% kcal for the lowest quartile) and the refined grains threshold (31% versus 35% - 38% in the lower quartiles) (**Figure 5b** and Supplemental Table 1).

There was no statistically significant interaction overall between IPR and race/ethnicity in relation to moderation food intake (F(4, 22) = 1.56, Prob > F = 0.22). However, the coefficient estimate for non-Hispanic Black adults was significantly more negative than that for Mexican American/Hispanic adults (**Table 2**). **Figure 6** shows that the estimated marginal slopes were negative for all groups, but the relationship was not statistically significant among Mexican Americans/Hispanic adults or those categorized as multi/other.

**Table 2.**
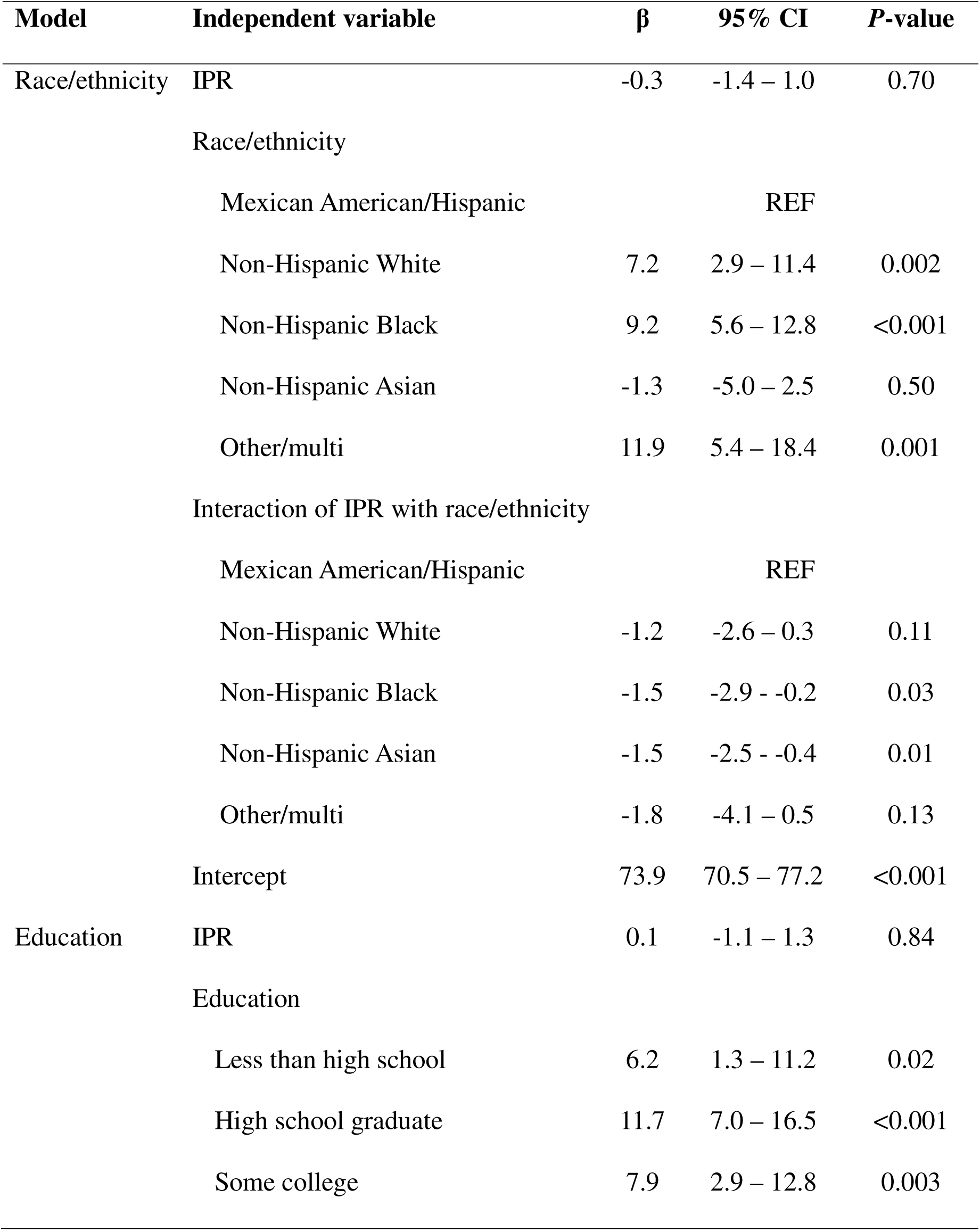
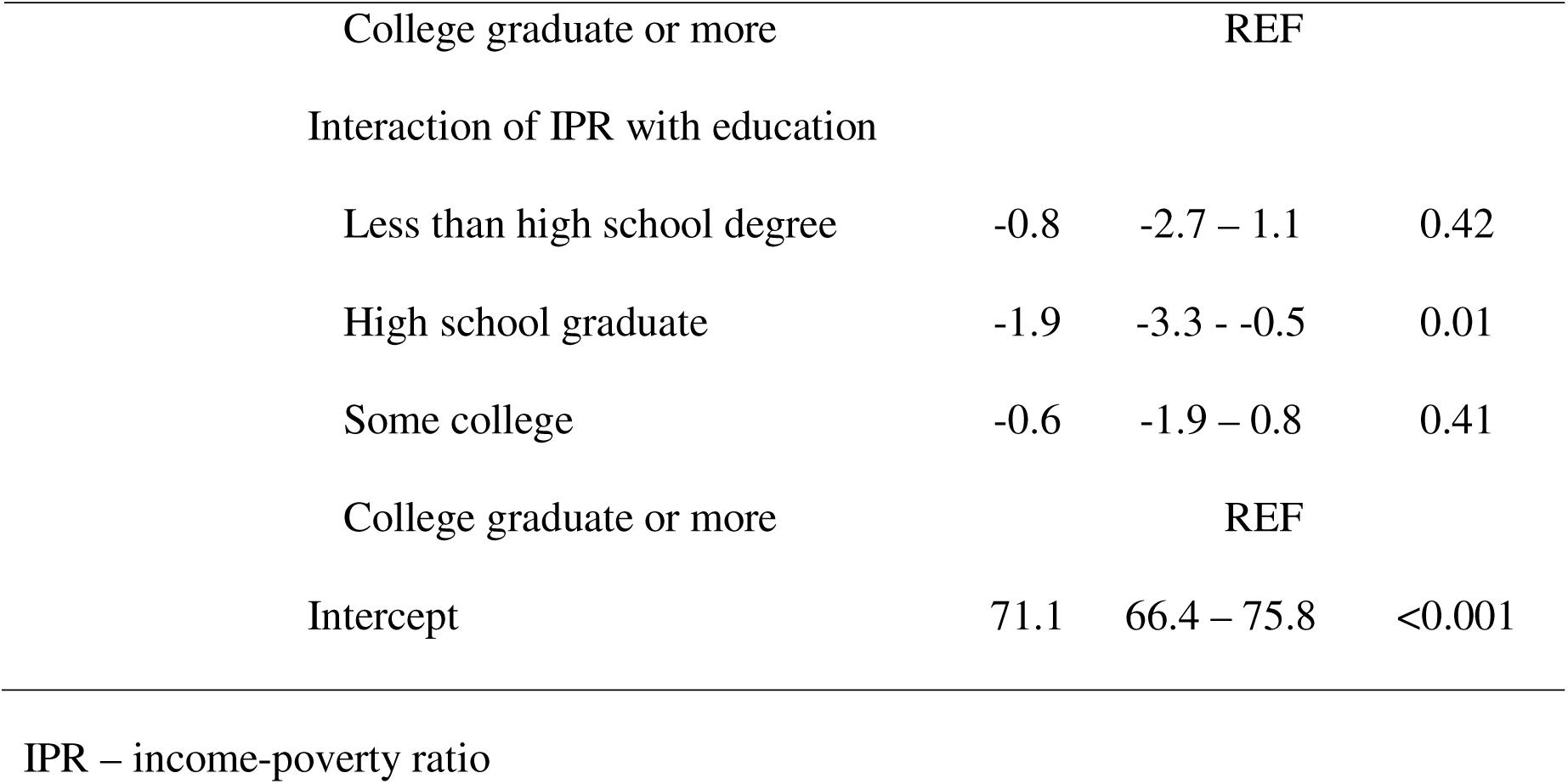
Coefficient estimates from regression models estimating the interactions of income poverty ratio (IPR) with race/ethnicity and education on moderation food intake (% kcal) in U.S. adults.

| Model | Independent variable | $\beta$ | 95% CI | P-value |
| --- | --- | --- | --- | --- |
| Race/ethnicity | IPR | -0.3 | -1.4 – 1.0 | 0.70 |
|  | Race/ethnicity |  |  |  |
|  | Mexican American/Hispanic |  | REF |  |
|  | Non-Hispanic White | 7.2 | 2.9 – 11.4 | 0.002 |
|  | Non-Hispanic Black | 9.2 | 5.6 – 12.8 | <0.001 |
|  | Non-Hispanic Asian | -1.3 | -5.0 – 2.5 | 0.50 |
|  | Other/multi | 11.9 | 5.4 – 18.4 | 0.001 |
|  | Interaction of IPR with race/ethnicity |  |  |  |
|  | Mexican American/Hispanic |  | REF |  |
|  | Non-Hispanic White | -1.2 | -2.6 – 0.3 | 0.11 |
|  | Non-Hispanic Black | -1.5 | -2.9 - -0.2 | 0.03 |
|  | Non-Hispanic Asian | -1.5 | -2.5 - -0.4 | 0.01 |
|  | Other/multi | -1.8 | -4.1 – 0.5 | 0.13 |
|  | Intercept | 73.9 | 70.5 – 77.2 | <0.001 |
| Education | IPR | 0.1 | -1.1 – 1.3 | 0.84 |
|  | Education |  |  |  |
|  | Less than high school | 6.2 | 1.3 – 11.2 | 0.02 |
|  | High school graduate | 11.7 | 7.0 – 16.5 | <0.001 |
|  | Some college | 7.9 | 2.9 – 12.8 | 0.003 |

|  |  |  |  |
| --- | --- | --- | --- |
| College graduate or more |  | REF |  |
| Interaction of IPR with education |  |  |  |
| Less than high school degree | -0.8 | -2.7 – 1.1 | 0.42 |
| High school graduate | -1.9 | -3.3 - -0.5 | 0.01 |
| Some college | -0.6 | -1.9 – 0.8 | 0.41 |
| College graduate or more |  | REF |  |
| Intercept | 71.1 | 66.4 – 75.8 | <0.001 |

IPR – income-poverty ratio

There was no statistically significant interaction between IPR and education in relation to moderation food intake overall (F(3, 23) = 2.6, Prob > F = 0.08) (**Table 2**). **Figure 7** shows a null association of income with moderation food intake in all education groups, except for an inverse association in high school graduates.

## Discussion

Mean moderation food intake ranged from 70% - 80% of energy intake across most segments of the U.S. adult population in 2017 – 2020. While some statistically significant differences were attributable to sex, age, income, race/ethnicity, and education, all subgroups obtained the majority of energy intake from moderation food, and the only subgroup with mean moderation food intake less than 70% kcal was non-Hispanic Asian adults (66%). Associations of income with moderation food intake did not vary by race or education, and moderation food intake differed more by race/ethnicity and education than by income. In the full sample, foods meeting thresholds for sodium, refined grains, and fat contributed more to total energy intake than foods meeting thresholds for added sugar or alcohol. Sociodemographic differences in overall moderation food intake were primarily due to differences in intakes of foods meeting thresholds for refined grains, sodium, and added sugar, with fewer differences in intakes of foods meeting thresholds for fat or alcohol.

These findings are consistent with previous studies that reported small but statistically significant socioeconomic disparities in adult diet quality^4,18,27–32^. In those studies, diet quality scores across all subgroups were far from optimal, with mean group differences ranging from <1% to 9%. Reports have often emphasized statistically significant differences while giving less attention to their small magnitudes, potentially contributing to the misconception that low diet quality affects only specific population subgroups. In contrast, our findings indicate that suboptimal dietary patterns are widespread, regardless of sex, income, race/ethnicity, or education, despite some statistically significant differences. While the DGA recommend that 85% of daily energy intake come from nutrient-dense foods, leaving only 15% for added sugars, fats, or alcohol ^1^, our findings suggest that dietary behaviors across all sociodemographic subgroups of U.S. adults are essentially reversed, with moderation food accounting for at least two-thirds of energy intake and non-moderation foods contributing only 20% to 30%. The magnitudes of subgroup differences were smallest by income-poverty ratio (1 – 4 points) and sex (3 points), and largest by age (8 points between the youngest and oldest adults) and race/ethnicity (7 – 13 points between non-Hispanic Asians versus other groups). The magnitude of the differences in intakes from foods meeting each moderation food component threshold were also generally less than 5 percentage points.

The consistently high moderation food intake observed across different sociodemographic subgroups suggests that sociodemographic factors may not be primary drivers of low diet quality in U.S. adults. This aligns with findings from the Consumer Expenditure Survey, which demonstrate that, across all income groups, the largest share of food-at-home spending (35%) goes to miscellaneous foods (e.g. frozen meals, snacks, canned and packaged soups, potato chips, condiments, seasonings), while 20% - 25% goes to meat, poultry, fish, and eggs; and 19% to fruits and vegetables^33^. These purchasing patterns suggest that households may prioritize spending on foods for convenience, social, cultural, emotional, or other purposes^34–36^, rather than adherence to dietary guidelines. Additionally, while food accessibility has been identified as a barrier to healthy eating for underserved groups, national data indicate that 6.1% of the U.S. population lives in areas with limited supermarket access, with 1.7% of households being both far from a food store and without vehicle access^37^.

The DGA’s focus on nutrients rather than foods to limit may have inadvertently weakened its effectiveness in discouraging intake of nutrients of concern in the U.S population. Since the 1970s, guidelines related to decreasing intakes have provided nutrient-based rather than food-based recommendations - a practice influenced by industry pressures^38^ and enabled by the closed nature of DGA meetings prior to the 2010 – 2015 cycle. Nutrient-based guidelines are more difficult for consumers to understand and implement than food-based recommendations^39^, partly since such guidelines require viewing and correctly interpreting nutrition labels that present nutrients per serving, which consumers must then accurately extrapolate to their total daily intake. Consequently, consumers may be unaware when intake of nutrients of concern exceeds recommended amounts. Although the shift to public DGA Committee meetings beginning with the 2010 – 2015 cycle has offered the opportunity to incorporate food-based recommendations for moderation components, the nutrient-based approach established during previous closed meetings has persisted. Another potential reason for the lack of current food-based guidelines to limit nutrients of concern may involve assertions, largely propagated by the food industry, that communicating health risks of specific foods inflicts psychological harm (e.g., “food shaming”)^40,41^. In contrast, empirical evidence shows that interpretive front-of-package labels improve consumer food choices without increasing stigma^42–44^. It will remain important to assess and monitor adverse psychological effects of any labels on at-risk groups, such as individuals with eating disorders, given limited evidence.

By contrast, the Chilean government adopted thresholds to classify packaged foods high in kcal, sugar, sodium, or saturated fat, and applied clear front-of-package labels along with restrictions on marketing to children and school sales. These policies significantly reduced consumer purchases and industry supply of high-sugar and high-sodium products, while also decreasing unhealthy marketing to children, limiting availability in schools, and improving consumers’ ability to identify and avoid unhealthy foods^45–50^. Collectively, this evidence demonstrates how policies that influence food environments can affect dietary behaviors, even though individual food choices are motivated by complex social, cultural, and emotional factors. Furthermore, it illustrates how regulatory approaches that use systematic food classification based on nutrient thresholds can drive meaningful changes in the food environment and serve as a powerful tool to improve diet quality and reduce chronic disease risk at the population level.

Several elements of this study strengthen the internal and external validity of the findings. The study used data from a large, nationally representative sample of the U.S. population that included administration of two dietary recalls using the Automated Multi-Pass Method. One limitation is that the use of self-reported dietary intake is susceptible to reporting bias that may weaken internal validity. However, we combined four years of NHANES data and used analytic methods to obtain reliable and generalizable estimates of overall intake of moderation food for multiple sociodemographic subgroups. We also examined potential confounding and effect modification between correlated characteristics (income, education, and race/ethnicity), which provides more valid estimates than bivariate analyses. As this was an observational study, it was descriptive rather than hypothesizing causal relationships.

## Conclusion

Moderation food intake is high in U.S. adults across multiple sociodemographic characteristics, indicating an urgent, population-wide need to replace moderation food with core foods that are aligned with nutritional requirements. Foods high in sodium, refined grains, and fat were the largest contributors to overall energy intake. The moderation food classification method may facilitate moving towards more explicit, food-based guidance and policy changes that modify the food environment to improve diet quality and chronic disease outcomes at the population level.

## Supporting information

Suuplemental Table 1

## Data Availability

All data are available online at: https://wwwn.cdc.gov/nchs/nhanes/continuousnhanes/default.aspx?Cycle=2017-2020

## Abbreviations

HEI: Healthy Eating Index
IPR: Income-Poverty Eatio
NHANES: National Health and Nutrition Examination Survey
NCHS: National Center for Health Statistics

## Acknowledgments

LL-designed research, analyzed data, wrote paper, had primary responsibility for final content; ACD – designed research, wrote paper; TRN – designed research, wrote paper. All authors have read and approved the manuscript.

## Declaration of generative AI and AI-assisted technologies in the writing process

During the preparation of this work, LL used ChatGPT [https://chat.openai.com/] and Perplexity [https://perplexity.ai]] to assist in improving the readability and clarity of this manuscript. After using these tools, LL reviewed and edited the content and takes full responsibility for the content of the publication.

