## Supplementary material for "Contribution of moderation foods to total energy intake by sociodemographic characteristics in U.S. adults: National Health and Nutrition Examination Survey, 2017 – 2020": Suuplemental Table 1

**Supplemental Table 1**. Mean percent intake from foods meeting each moderation food component threshold^1^ overall and by sociodemographic characteristics in U.S. adults.^2^

|  | **Added sugar** | | **Sodium** | | **Refined grains** | | **Fat** | | **Alcohol** | |
| --- | --- | --- | --- | --- | --- | --- | --- | --- | --- | --- |
|  | Mean  % kcal | 95%CI | Mean  % kcal | 95%CI | Mean  % kcal | 95%CI | Mean % kcal | 95%CI | Mean  % kcal | 95%CI |
| **Full sample** | 19 | 18 – 20 | 31 | 30 – 33 | 35 | 34 – 36 | 35 | 34 – 36 | 4 | 3 – 4 |
| **Sex** |  |  |  |  |  |  |  |  |  |  |
| Female | 20 | 19 – 21 | 29 | 28 – 31 | 34 | 32 – 37 | 28 | 26 – 29 | 3 | 2 – 3 |
| Male | 18 | 17 – 19 | 33 | 32 – 35 | 36 | 35 – 37 | 27 | 26 – 28 | 5 | 4 – 5 |
| **Age, years** |  |  |  |  |  |  |  |  |  |  |
| 20 – 39 | 19 | 17 – 20 | 36 | 34 -38 | 38 | 37 – 39 | 26 | 25 – 27 | 4 | 3 – 5 |
| 40 – 59 | 20 | 19 – 21 | 30 | 29 – 31 | 34 | 33 – 35 | 27 | 26 – 28 | 4 | 4 – 5 |
| ≥ 60 | 19 | 18 - 20 | 27 | 26 - 28 | 32 | 30 – 33 | 30 | 28 – 32 | 3 | 2 – 4 |
| **Race/ethnicity** |  |  |  |  |  |  |  |  |  |  |
| Mexican American/Hispanic | 17 | 16 – 18 | 32 | 31 – 34 | 34 | 32 – 36 | 24 | 23 – 25 | 3 | 2 – 3 |
| Non-Hispanic white | 20 | 19 – 21 | 31 | 29 – 33 | 34 | 33 – 35 | 29 | 28 – 31 | 4 | 4 – 5 |
| Non-Hispanic black | 20 | 19 – 21 | 34 | 32 – 35 | 39 | 37 – 40 | 27 | 26 – 28 | 4 | 3 – 4 |
| Non-Hispanic Asian | 13 | 12 – 15 | 27 | 26 – 29 | 35 | 33 – 37 | 18 | 16 – 20 | 2 | 1 – 2 |
| Other/multi | 22 | 19 - 25 | 31 | 27 - 36 | 37 | 34 - 40 | 28 | 25 - 32 | 4 | 2 - 6 |
| **Education** |  |  |  |  |  |  |  |  |  |  |
| < High school graduate | 20 | 18 – 21 | 30 | 28 – 31 | 36 | 34 – 37 | 26 | 25 – 28 | 3 | 2 – 3 |
| High school graduate | 21 | 19 – 22 | 33 | 31 – 35 | 38 | 37 – 40 | 28 | 27 – 29 | 3 | 3 – 4 |
| Some college | 20 | 19 – 21 | 32 | 31 – 34 | 35 | 34 – 37 | 28 | 27 – 29 | 4 | 3 – 4 |
| ≥ College graduate | 17 | 16 - 18 | 30 | 28 - 31 | 31 | 30 - 33 | 27 | 26 – 29 | 4 | 4 - 5 |
| **IPR quartile** |  |  |  |  |  |  |  |  |  |  |
| 1 (lowest income) | 22 | 20 – 23 | 32 | 30 -33 | 37 | 36 – 38 | 27 | 25 – 28 | 3 | 2 – 3 |
| 2 | 20 | 19 – 21 | 31 | 29 – 32 | 36 | 35 – 38 | 27 | 25 – 28 | 3 | 2 – 4 |
| 3 | 20 | 18 – 21 | 33 | 31 – 35 | 36 | 34 – 37 | 29 | 28 – 31 | 3 | 3 – 4 |
| 4 (highest income) | 17 | 16 - 19 | 31 | 28 - 33 | 33 | 31 – 35 | 28 | 27 – 29 | 5 | 4 - 6 |

^1^Moderation food component thresholds included: added sugar >20% energy, sodium >460mg per serving, refined grains >50% of total grains or >10:1 ratio of carbohydrate to fiber content, saturated fat >20% energy, total fat >9% by weight (applied only to vegetables, sweets, and snacks), and all alcoholic beverages.

^2^Differences of at least 5 percentage points are highlighted. Yellow cells indicate that a group mean is 5 or more percentage points lower than another group mean. Red cells indicate that a group mean is 5 or more percentage points higher than another groups. Orange cells indicate that a group mean is both 5 or more percentage points lower than another group and 5 or more percentage points higher than another group.
